# ESTIMATION OF COVID-19 CASES IN FRANCE AND IN DIFFERENT COUNTRIES: HOMOGENEISATION BASED ON MORTALITY

**DOI:** 10.1101/2020.04.07.20055913

**Authors:** Marc Dhenain

**Affiliations:** Académie Vétérinaire de France, 34, rue Bréguet, 75011 Paris, France; Académie Nationale de Médecine, 16 rue Bonaparte, 75006 Paris, France; Centre National de la Recherche Scientifique (CNRS), Université Paris-Sud, Université Paris-Saclay UMR 9199, Laboratoire des Maladies Neurodégénératives, 18 Route du Panorama, F-92265 Fontenay-aux-Roses, France; Commissariat à I’Energie Atomique et aux Energies Alternatives (CEA), Institut François Jacob, Molecular Imaging Research Center (MIRCen), 18 Route du Panorama, F-92265 Fontenay-aux-Roses, France

**Keywords:** Covid-19, Estimated number of cases, Mortality, Prevalence

## Abstract

Every day the authorities of different countries provide an estimate of the number of persons affected by Covid-19 and a count of fatality. We propose to use the fatality reported in each country to provide a better estimate (C_t0-estimated_) of the number of cases at a given time t_0_.

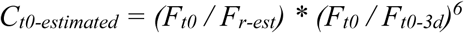

With Ft_0_: number of fatalities reported in a country at time t_0_; F_t0_: number of fatalities reported in a country at time t_0_ minus 3 days; F_r-est_: estimated fatality rate. Based on C_t0-estimated_ calculated using a fatality rate of 2%, we assessed the number of cases April 10^th^, 2020 in Belgium, China, France, Germany, Iran, Italy, South Korea, Netherlands, Spain, United Kingdom and USA. This number reached 2,872,097 in France and 924,892 persons in Germany. This work suggests a very strong underestimation of the number of cases of people affected, with a notification index often lower than 5%. The proposed formulas also make it possible to evaluate the impact of policies to prevent the spread of epidemic.

## INTRODUCTION

The Sars-CoV-2 coronavirus infection that causes Covid-19 has spread worldwide leading to significant deaths (European Center for Disease Prevention and Control 2020). Every day authorities of different countries provide an estimate of the number of affected persons and a count of fatalities (Dong et al. 2020, https://ourworldindata.org/covid-testing, https://github.com/CSSEGISandData/COVID-19/tree/master/csse_covid_19_data/csse_covid_19_time_series). Knowing the number of affected subjects is critical for implementing strategies to protect populations and for ending the crisis. Figures reported by different countries reveal strong differences and only partly reflects the reality (Table I). For example, the day their death toll approached 3,000 people, France had 44,550 people affected versus 80,537 for China and 122,171 for Germany. Calculating the case fatality rate (F_r_) on a given day (t_0_) is another way to objectify differences between countries. At first sight,

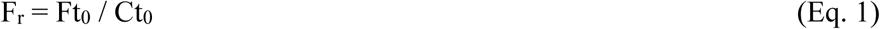

**Table I:** Fatality rates in different countries when the number of deaths approached 3,000 people (or the last figure available when the 3,000 deaths were not reached April 10^th^ 2020) (https://github.com/CSSEGISandData/COVID-19/tree/master/csse_covid_19_data/csse_covid_19_time_series)

| Country | Day<br>($t_0$) | Fatalities<br>at $t_0$<br>($F_{t_0}$) | Cases<br>at $t_0$<br>($C_{t_0}$) | Fatality<br>rate<br>$F_r$ | Cases<br>at $t_0-18j$<br>($C_{t_0-18d}$) | Fatality<br>rate<br>$F_{r-18d}$ |
| --- | --- | --- | --- | --- | --- | --- |
| Belgium | April 10 | 3 019 | 26 667 | 11.3% | 3743 | 80.7% |
| China | March 5 | 3 015 | 80 537 | 3.7% | 70 513 | 4.3% |
| France | March 30 | 3 024 | 44 550 | 6.8% | 2 281 | 132.6% |
| Germany | April 10 | 2 767 | 122 171 | 2.3% | 29 056 | 9.5% |
| Iran | April 1 | 3 036 | 47 593 | 6.4% | 12 729 | 23.9% |
| Italy | March 18 | 2 978 | 35 713 | 8.3% | 1 128 | 264.0% |
| South-Korea | April 10 | 208 | 10 450 | 2.0% | 8 961 | 2.3% |
| Netherlands | April 10 | 2 511 | 23 097 | 10.9% | 4 749 | 52.9% |
| Spain | March 24 | 2 808 | 39 885 | 7.0% | 400 | 702.0% |
| United Kingdom | April 2 | 2 921 | 33 718 | 8.6% | 1 140 | 256.2% |
| USA | March 30 | 2 978 | 161 807 | 1.8% | 1 163 | 256.1% |

With Ft_0_ = number of fatalities reported on day t_0_; Ct_0_ = number of cases reported on day t_0_. The day (the closest to April 10^th^, date of redaction of this article) when the death toll of different countries was the closest to 3,000 people, three countries (Germany, South Korea, and the United States) had fatality rates close to 2%; seven countries (Belgium, France, Iran, Italy, the Netherlands, Spain, and the United Kingdom) had rates between 6% and 12%, and China had an intermediate value of 3.7% (Table I).

Patients who die on any given day were infected much earlier, and thus the denominator of the fatality rate should be the total number of patients infected at the same time as those who died (Baud et al. 2020). This is particularly true as the rates of evolution of the pandemic evolve differently in various countries: in March 2020, the number of people affected increased sharply from day to day in France, while it was stable in China.

A better estimate of fatality rate is thus:

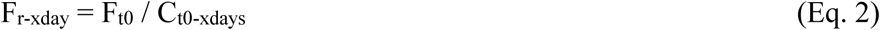

With Ct_0-xdays_ = number of cases reported on day t_0_ minus x days, with x = average time-period from onset of symptoms to death.

An average duration of 18 days is reported between the onset of symptoms and the death of Covid-19 patients (Ruan et al. 2020; Verity et al. 2020; Zhou et al. 2020). Thus, the adjusted Fatality rate (F_r-18d_) that takes into account this average delay is (Flaxman et al. 2020).

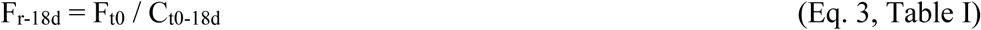

With C_t0-18d_ = number of cases reported on day t_0_ minus 18 days. The calculation of F_r-18d_ reveals widening gaps between countries compared to F_r_ with variations ranging from 2.3% (South Korea) to more than 700% for Spain.

When comparing F_t0_ and C_t0-18d_ in different countries (Fig. 1), we see a linear relationship between mortality at t_0_ and the number of cases at t_0-18days_ for all countries (Pearson linear correlation test, p<0.05 except for Belgium (p=0.07) due to the small number of points (n=3)). The slopes of the regression lines fitting the data vary widely between countries, which is consistent with variable F_r-18_. The values of fatality rate F_r-18_ based on the cases reported by the different countries are therefore unreliable, in part because the number of cases reported in different countries is not reliable (different testing strategies in different countries).

**Figure 1:**
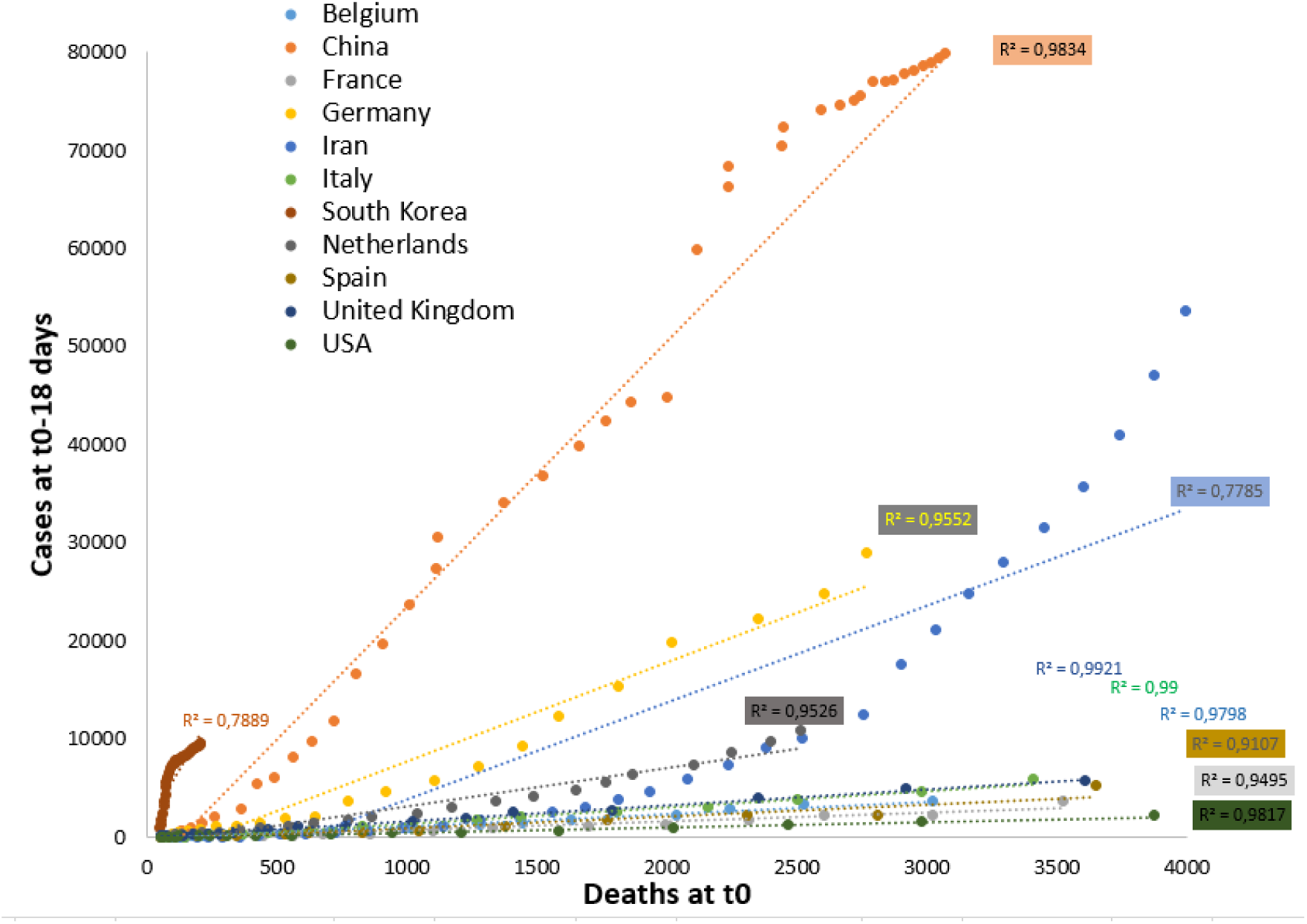
Relationships between deaths a given day (t0) and the number of cases eighteen days before (t_0-18days_) in different countries. The figure includes only values between 50 and 4000 deaths (or less if the number of deaths was lower in the country April 10^th^).

How to assess more precisely the number of people affected using a similar method for different countries? We offer a simple method using the number of deaths reported by each country to estimate and compare the rate of people affected by Covid-19. This method relies on three first assumptions: 1. The number of deaths reported by each country is reliable; 2. The fatality rate (F_r_) is known and similar in different countries; 3. The average time between the onset of symptoms and death is known (here considered 18 days). Based on these assumptions, one can calculate the number of cases presented eighteen days before a given day (t_0_). Two methods are then proposed to infer the number of cases, eighteen days later, at time t_0_. The first one relies on the time-dependent increase in the number of cases reported in databases during these 18 days. The second one models the evolution of the number of cases based on daily rate of changes of the number of estimated cases 18 days before t_0_.

## METHODS

### Estimation of the number of cases at time t0 minus 18 days based on fatality rate

One way to estimate the number of Covid-19 cases is to infer the number of cases based on the number of death and the fatality rate calculated from well-controlled studies using the following formula

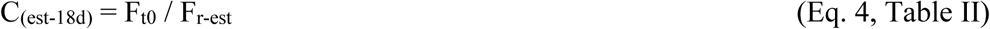

With C_(est-18d)_: number of cases estimated 18 days before t_0_; Ft_0_ = number of deaths reported on day t_0_; F_r-est_ = estimated fatality rate from well-controlled studies. F_r-est_ can be assessed from well-controlled studies based on residents of mainland China, travelers returning from mainland China, repatriated from China, passengers on the *Diamond-Princess* cruise ship (values of 0.7 to 3.6% (Verity et al. 2020)). Here, based on this last study, we proposed to use F_r-est_ = 2%.

Knowing C_(est-18d)_ eighteen days before t_0_, one needs to assess the progression rate of the cases during the 18 last days to estimate the number of cases at day t_0_ (C_t0-estimated)_. We tested two methods to assess this progression.

### Estimation of progression of cases for 18 days based on reported number of cases

One can assume that progression of estimated cases (P_18d_) reflects the time-dependent increase in the reported number of cases during the same time-period. In that case,

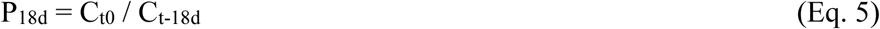

With C_t0_: number of cases reported in a country at time t_0_; C_t-18d_: number of cases reported in the country at time t0 minus 18 days. Thus,

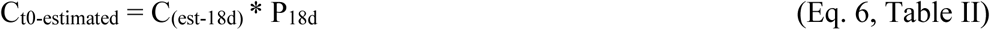

### Estimation of progression of cases for 18 days based on 3-day rate of change of the estimated C(_est-18d_)

Another option to assess the progression of cases for 18 days is to calculate the daily rate of change of the number of estimated cases (R_d_) or alternatively 3-day rate of change (R_3d_) of the number of estimated cases

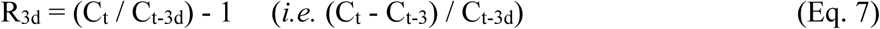

With C_t0_: number of cases reported at time t_0_; C_t0-3d_: number of cases reported at time t_0_ minus 3 days.

The last day when this calculation is possible is 18 days before t_0_.

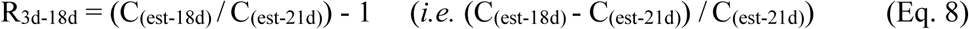

With C_(est-18d)_: estimated cases 18 days before t_0_ (see Eq. 4); C_(est-21d)_: estimated cases three days before. Assuming that the progression of the estimated cases follows an exponential model then

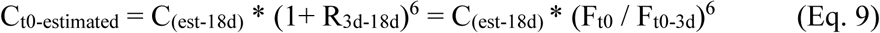

With F_t0-3d_: number of fatalities reported in a country at time t_0_ minus 3 days. The exponent 6 represents the period of the model as 6 * 3 days = 18 days. This estimation supposes that R_3d_ does not evolve with time, during the last 18 days.

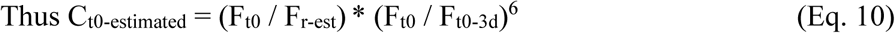

### Comparison with basic reproductive potential of the pathogen (R_0_)

The basic reproductive potential of the pathogen (R_0_) is an important index in epidemiology. It is defined as the average number of secondary cases arising from a primary case in an entirely susceptible population. Another critical parameter is the mean generation time (Tg) *i.e*., the time between the infection of a primary case and the infection of a secondary case (Flaxman et al. 2020; Keeling & Rohani, 2008).

Assuming that the progression of Covid-19 follows a SEIR model based on four compartments (Susceptible, Exposed, Infectious, and Recovered subjects (Keeling & Rohani, 2008)), the increase in prevalence during the invasion phase of the disease is estimated as

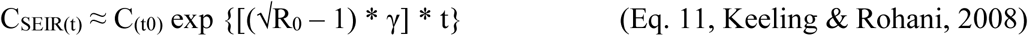

With C: number of infected cases at a time t or t_0_; R_0_: the basic reproductive potential of the pathogen; *γ*: the recovery rate *γ* with 1/ *γ*: the infectious period (Di).

This equation allows to estimate the doubling time for cases (T2) if C_SEIR(t)_/C_(t0)_=2.

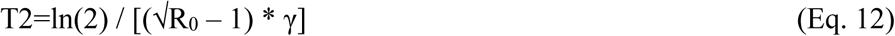

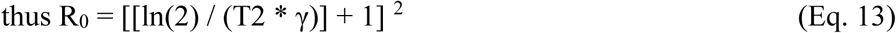

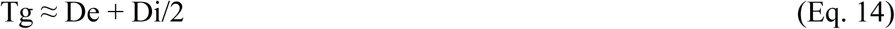

if Covid-19 is modeled with a SEIR model (with De = exposition time during which a subject is exposed but not infectious), and De ≈ Di (Li et al., 2020).

One can thus estimate that

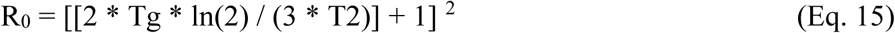

Using the daily rate of change (R_d_) (or R_3d_ / 3), and the “rule of 70”, one can estimate

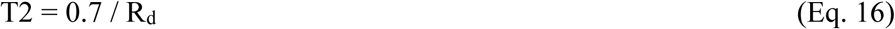

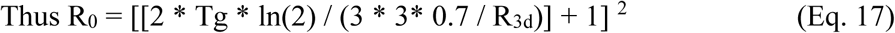

Tg can be estimated to be 6.5 according to (Flaxman et al. 2020).

## RESULTS

### Estimations relying on number of cases at time to minus 18 days

Using the (Eq. 4) and international databases (Dong et al. 2020, https://github.com/CSSEGISandData/COVID-19/tree/master/csse_covid_19_data/csse_covid_19_time_series), we estimated the number of people that have been affected by Covid-19 eighteen days before April 10^st^ 2020 in different countries (Table II). This estimation was 659,850 persons in France and 138,350 in Germany. We then proposed two different methods to infer the number of cases 18 days latter (Fig. 2). Method based on P_18d_ evaluation using the reported number of cases during the same time-period suggested a time-dependent decrease of number of cases in some countries (e.g. Germany (Fig. 2C) or USA (Fig. 2D)), which is not consistent. Method based on the evaluation of R_3d_ provided a better correspondence with the estimated cases in all tested countries (Fig. 2). We thus retained results from the R_3d_ method for further analyses. Estimation of the number of cases based, April 10^th^ was 2,872,097 in France, 924,892 in Germany, 1,811,469 in Spain, 4,240,198 in the United Kingdom and 9,035,229 in the United States (Table II, Fig. 2, Fig. 3A). R_3d_ evaluation can further be used to assess the basic reproductive potential of the pathogen (R_0_) based on (Eq. 17). We reported R_0_ values from 1.0 (China) to 2.86 (Belgium) (Table II).

**Figure 2:**
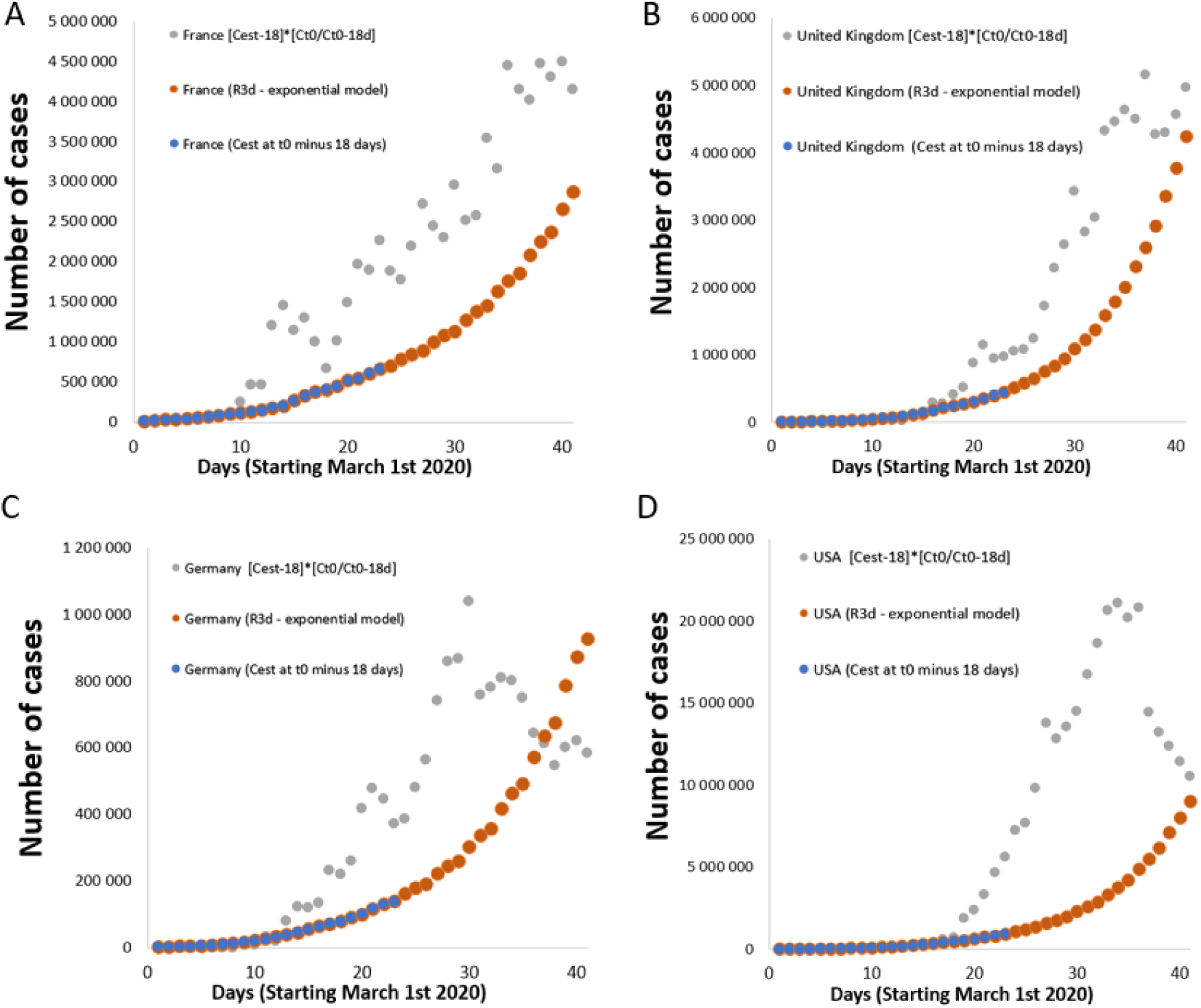
Comparison of evolution of estimated Covid-19 cases in France (A), United Kingdom (B), Germany (C), and USA (D) from March 1^st^ 2020 to April 10^th^ 2020. The estimated values corresponding to the number of cases estimated 18 days before t_0_(C_est-18d_), based on the number of deaths at t0 are displayed in blue. The progression of these cases, taking into account a multiplying factor (P_18d_) which reflects the increase over time in the number of cases declared during the same period is noted in gray. This calculation method leads to a large number of cases and to daily variations. The orange marks correspond to a model based on R3d ((1 + C_est-3d_) * R_3d_). It provides curves which follow the values corresponding to C_est_ based on fatalities estimated 18 days before a given day (blue marks).

**Fig. 3.**
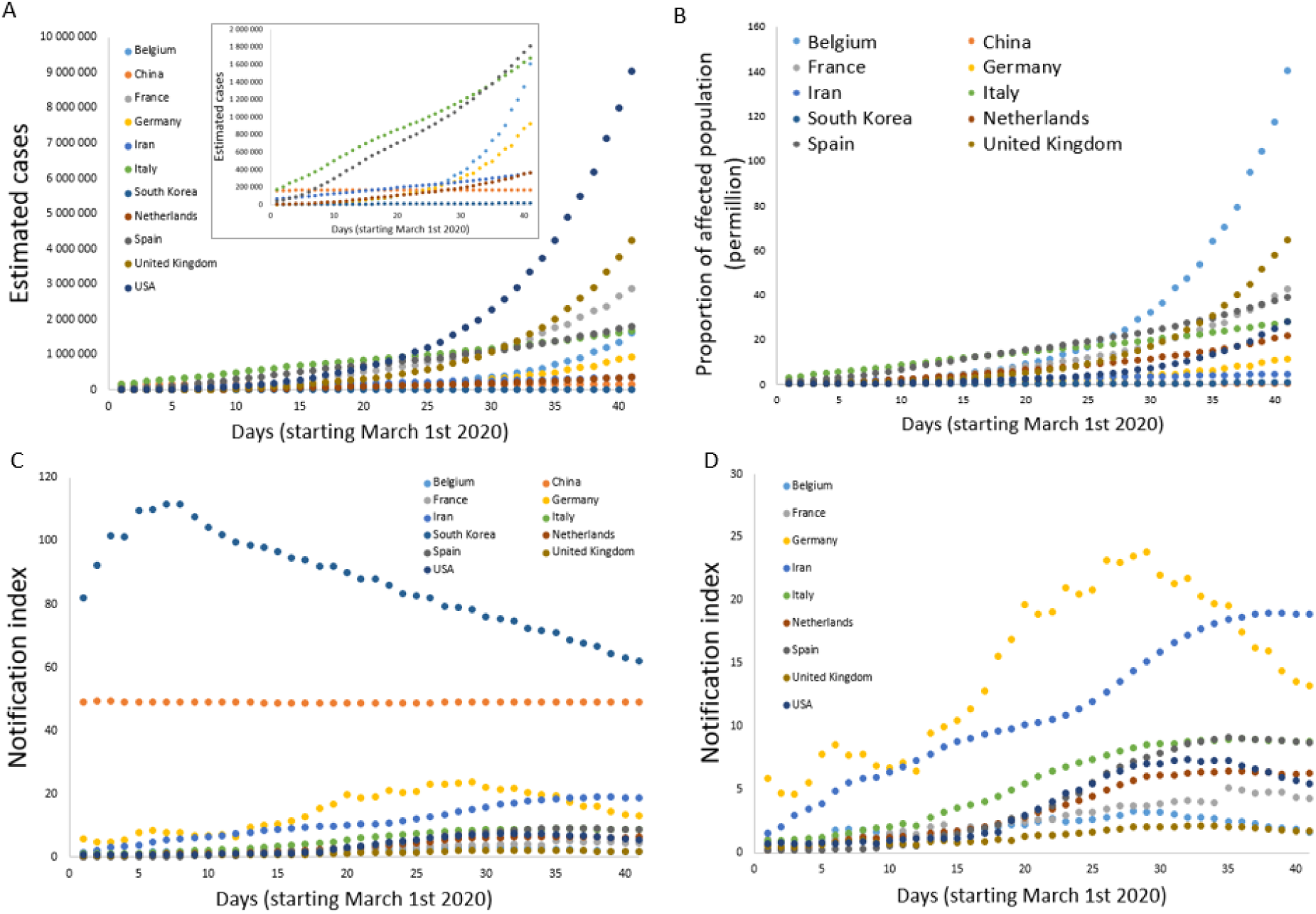
Comparison of estimated cases and related parameters in different countries. A. estimated cases in different countries. B. Proportion of affected person compared to the country population. C-D. Notifications indexes reflecting the number of cases reported by different countries compared to the estimated number of cases (percentages).

**Table II:** Estimation of the number of cases in different countries April 10^st^ (t_0_) using different methods and an estimated Fatality rate (F_r-est_) of 2%. Numbers of cases estimated with different methods are provided using delays of 18 or 12 days between symptom occurrence and death.

|  |  |  |  |  | Method 1&2 | Method 1<br>Progression rate from actual cases |  | Method 2<br>Estimation from 3-days rate of change |  |  |  |  |  |
| --- | --- | --- | --- | --- | --- | --- | --- | --- | --- | --- | --- | --- | --- |
| Country | Population (million) | Reported cases at $t_0$<br>(April 10)<br>($C_{t_0}$) | Reported deaths at $t_0$<br>(April 10)<br>($F_{t_0}$) | Reported cases at $t_{0-18d}$<br>(March 23)<br>($C_{t_0-18d}$) | Estimated cases at $t_{0-18d}$<br>(March 23)<br>($C_{est-18d}$) | Progression rate from $t_{-18}$ to $t_0$<br>$P_{18d} = C_{t_0} / C_{t_0-18d}$<br>(April 10)<br>$P_{18d}$ | Estimated Cases<br>(April 10)<br>$C_{t_0-estimated}$ | $R_{3d-18j}$<br>(April 10)<br>$C_{t_0-estimated}$ | $R_0$<br>(April 10)<br>$C_{t_0-estimated}$ | Estimated cases<br>(April 10)<br>$C_{t_0-estimated}$ | Estimated cases<br>(April 10)<br>$C_{t_0-estimated}$ | Estimated cases<br>(April 10)<br>$C_{t_0-estimated}$ | Estimated cases<br>(April 10)<br>$C_{t_0-estimated}$ |
| Delay | | | | | | | $t_{0-18d}$ | $t_{0-18d}$ | $t_{0-18d}$ | $t_{0-18d}$ | $t_{0-18d}$ | $t_{0-18d}$ | $t_{0-12d}$ |
| $M_{r-est}$ | | | | | | | 2% | 2% | 2% | 2% | 4% | 15% | 2% |
| Belgium | 11,476 | 26 667 | 3019 | 3 743 | 150 950 | 7,12 | 1 075 443 | 0,48 | 2,86 | 1 609 257 | 804 628 | 214 568 | 683 894 |
| China | 1,384,688 | 82 941 | 3340 | 81 498 | 167 000 | 1,02 | 169 957 | 0,00 | 1,00 | 168 508 | 84 254 | 22 468 | 168 003 |
| France | 67,795 | 124 869 | 13197 | 19 856 | 659 850 | 6,29 | 4 149 618 | 0,28 | 1,95 | 2 872 097 | 1 436 049 | 382 946 | 1 645 302 |
| Germany | 83,073 | 122 171 | 2767 | 29 056 | 138 350 | 4,20 | 581 717 | 0,37 | 2,35 | 924 892 | 462 446 | 123 319 | 482 669 |
| Iran | 82,022 | 68 192 | 4232 | 23 049 | 211 600 | 2,96 | 626 033 | 0,09 | 1,28 | 360 726 | 180 363 | 48 097 | 294 241 |
| Italy | 60,360 | 147 577 | 18849 | 63 927 | 942 450 | 2,31 | 2 175 668 | 0,10 | 1,31 | 1 674 559 | 837 279 | 223 275 | 1 364 771 |
| South-Korea | 51,709 | 10 450 | 208 | 8 961 | 10 400 | 1,17 | 12 128 | 0,08 | 1,25 | 16 811 | 8 406 | 2 242 | 14 306 |
| Netherlands | 17,282 | 23 097 | 2511 | 4 749 | 125 550 | 4,86 | 610 619 | 0,20 | 1,64 | 365 882 | 182 941 | 48 784 | 240 570 |
| Spain | 46,935 | 158 273 | 16081 | 35 136 | 804 050 | 4,50 | 3 621 909 | 0,14 | 1,46 | 1 811 469 | 905 735 | 241 529 | 1 345 788 |
| United Kingdom | 65,761 | 73 758 | 8958 | 6 650 | 447 900 | 11,09 | 4 967 851 | 0,45 | 2,72 | 4 240 198 | 2 120 099 | 565 360 | 2 041 524 |
| USA | 328,240 | 496 535 | 18586 | 43 847 | 929 300 | 11,32 | 10 523 638 | 0,46 | 2,75 | 9 035 229 | 4 517 615 | 1 204 697 | 4 086 903 |

This analysis is based on an estimated fatality rate from well-controlled studies (M_r-est_) of 2%. The estimated number of cases must be halved if the mortality rate used jumps from 2 to 4% (Table II). It must be doubled if the fatality rate used drops from 2 to 1%. Some authors suggest that the real fatality rate for Covid-19 could be 5.6 to 15.6% (Baud et al. 2020). If the calculation uses a fatality rate of 15%, then the estimated number of cases drops to 382,946 for France, but it becomes lower than the number of cases actually reported for some countries (e.g. 2,242 *versus* 10,450 for South Korea), which is not consistent (Table II).

In our study, we set the delay between the onset of symptoms and death at 18 days based on robust data from the literature (Ruan et al. 2020; Verity et al. 2020; Zhou et al. 2020) and delays used in other models (Flaxman et al. 2020). Lowering this delay, for example to 12 days, sharply decreases the number of estimated cases (*e.g*. 1,645,302 for France (Table II)) although it remains high compared to figures reported by most countries.

Using estimations based on R_3d_ model, with delay of 18 days between the onset of symptoms and death and fatality rate of 2%, we could thus compare estimated cases of Covid-19 in different countries (Fig. 3A), proportion of cases in different countries (Fig. 3B), as well as notification indexes which is the ability to report cases (Fig. 3C-D). These data highlight strong discrepancies between countries. It suggests a high proportion of affected persons in Belgium. It also shows notification indexes that varies from 60 to 80% in Korea while it is below 5% in most countries.

### Comparison of estimated cases with the number of cases reported afterwards

The number of cases evaluated between March 16 and April 10, 2020 from the number of cases at time t_0_ minus 18 days and the R_3d_ model was compared with cases estimated from the number of cases at time t_0_ minus 18 days (without the R_3d_-based model) calculated from mortality data collected between April 11 and 28, 2020 (Fig. 4). The cases estimated with the R_3d_ model were higher than those estimated a posteriori. This can easily be explained by the reduction in the spread of the disease (and thus of R_3d_) in the past 18 days following the containment measures in many countries. The three-day rate of change in the number of estimated cases (R_3d_) we used is overstated.

**Fig. 4.**
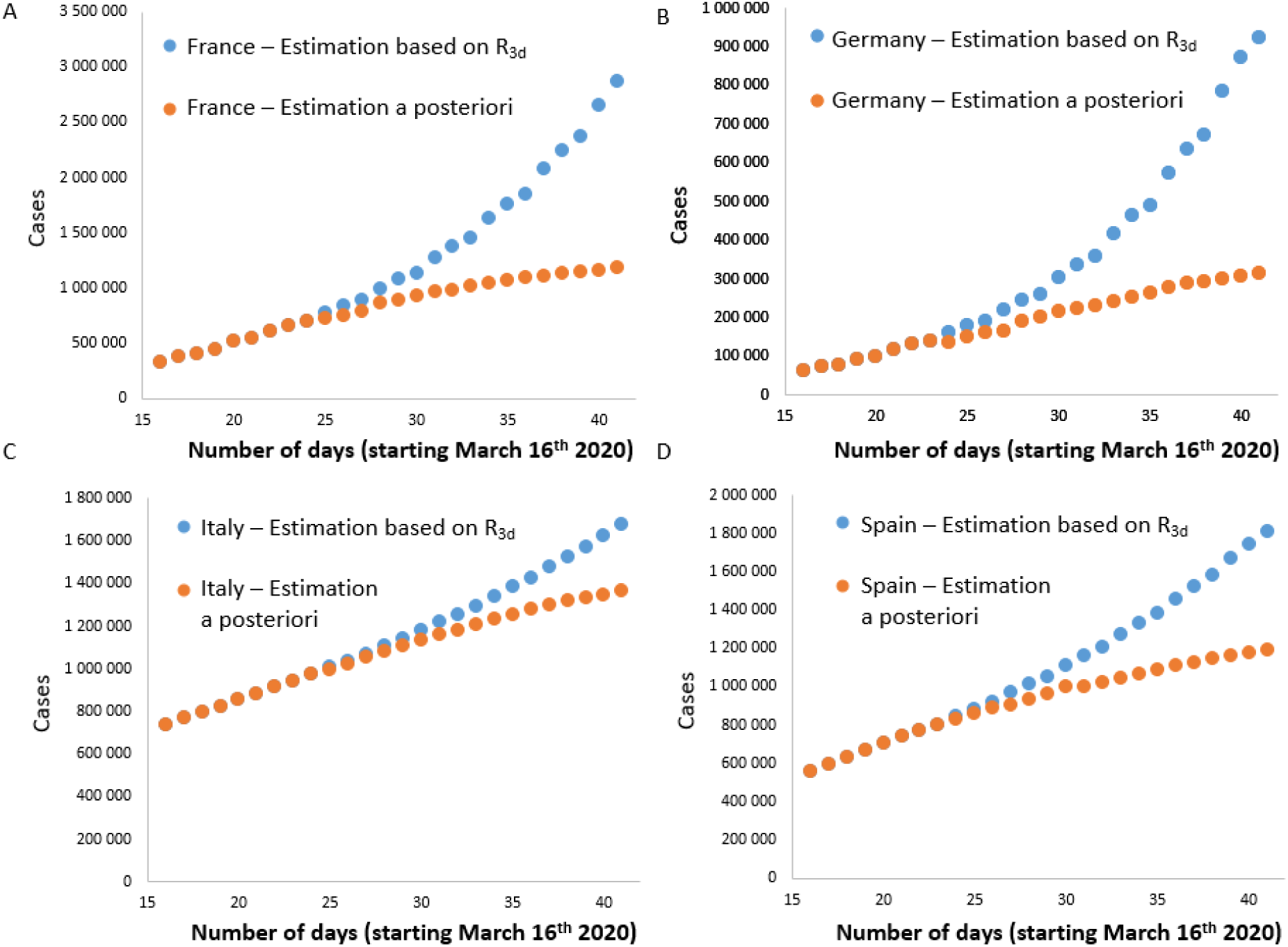
Comparison of cases estimated in different countries from day 16 (March 16^th^) using a method based on fatality rates reported from March 16 to April 10 with an estimate based on R_3d_ (blue). Results of an a posteriori method following the fatality rates reported from April 3 to April 28 are shown in orange.

The measurements that we have made allow to assess the impact of containment policies. For example, for France, the cases estimated on April 10 were based on an R_3d_ of 0.28 (measured on March 23). Five days before this date (March 19), R_3d_ was 0.50. Using an R_3d_ of 0.50 (instead of 0.28) leads to an estimate of 7,516,104 cases on April 10. The reduction of R_3d_ from 0.50 to 0.28 thanks to containment therefore prevented the appearance of 4,644,007 new cases in France (7,514,104 - 2,872,097). The number of cases estimated from the deaths which occurred on April 28, 2020 was in fact 1,141,450. Thus, the number of cases actually avoided is 7,514,104 - 1,181,450 = 6,332,654. This represents 126 653 avoided deaths.

## DISCUSSION

It is essential to assess the number of persons affected by Covid-19 in all countries of the world to stem this crisis. We propose to use the mortality reported by each country at a time t_0_ to create an index of the number of real cases at this same time t_0_. Mortality at t_0_ makes it possible to estimate the number of cases 18 days earlier (C_(est-18d)_). Then, the rate of change over time of the estimated cases (C_(est)_) is evaluated over 3 days (R_3d_). This rate is used to modulate C_(est-18d)_, and calculate the number of cases at time t0. This calculation leads to the following equation

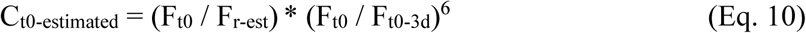

With F_t0_: number of fatalities reported in a country at time t_0_; F_t0_: number of fatalities reported in a country at time t_0_ minus 3 days; F_r-est_: estimated fatality rate. This analysis is based on four assumptions: 1. The number of deaths reported by each country is reliable, 2. The estimated fatality rate among people affected is known (F_r-est_, here considered as 2%), 3. The average time between the onset of symptoms and death is known (here considered 18 days). 4. The rate of variation over three days of the estimated cases (R_3d_) does not change during the last 18 days. This last condition is not entirely exact because, thanks to containment policies, the rate of change over three days decreased continuously until the beginning of May 2020. C_t0-estimated_ is thus overestimated as shown by the comparison of the values obtained with measurements a posteriori, that is to say without taking into account the modeling of the last 18 days. Estimating the 3-day rate of change in the number of estimated cases (C_(est-18d)_ - Eq.4) can be used to assess R_0_. The values of R_0_ that we have reported (from 1.0 (China) to 2.86 (Belgium)) are consistent with data from the literature (for example R_0_ = 4.0 in Flaxman et al. 2020, at the beginning of pandemic). Conversely, the estimation of R_0_ on the basis of an epidemiological model (cf. for example (Flaxman et al. 2020)) could be used to calculate R_3d_ (cf. Eq. 17) and refine the estimation of the number of cases. It would also be possible to smooth out the risks of daily variations in the calculation of R_3d_ by using average measures of R_3d_ over a longer period of time.

Our analyses showed that the number of Covid-19 cases in several country greatly exceeds the number of cases presented in international databases (2,872,097 (or 1,181,450 cases with a posteriori measures) versus 124,869 for France on April 10^th^, 2020). The very high values of estimated cases that we report are consistent with those evaluated with another method by (Flaxman et al. 2020). For example, we report 1.8 million cases in Spain while Flaxman reports 7.0 million on March 28^th^. Our calculation relies on a relatively simple method while that of Flaxman uses more complex analyzes (hierarchical semi-mechanistic Bayesian model). Our model used a fatality rate of 2% while several strongly controlled international studies reported rates of 0.7 to 3.6% (Verity et al. 2020). Values from 0.5 to 4% could thus be other reasonable options to estimate fatality rate. One of the limitations of our model is that fatality rates can change from one country to another, for example depending on the distribution of the population of different age groups that have different susceptibility to Covid-19. Also, it is possible that death rate changes over time in a given country, for example because of the saturation of hospitals or the correction of mortality figures to include non-counted cases (as done in France between April 1 and 4, 2020 to include mortality in nursing homes). We fixed a single value for the time between symptom occurrence and death (18 days). In reality, this time is variable with a 95% credible interval of 16.9 to 19.2 or more (Verity et al. 2020). We however considered that using such interval would make the model more complicated without strongly adding reliability compared to other potential sources of errors. Our analysis is based solely on the number of people who have died with confirmed Covid-19 cases. It is therefore essential that all countries are able to provide very reliable Covid-19 death values. Finally, note that to know the number of actual cases in a country at a given time, we must subtract from the estimates presented here the number of people healed, including those whose disease has not been identified.

To conclude, our model questions the small number of people reported to be affected by Covid-19 in most countries compared to the large numbers we estimate. This difference could be explained by a large underestimation of the “mortality rate”. For example, in France, the “estimated mortality rate” should be changed from 2 to ~ 46% to decrease the estimate of the number of cases from 2,872,097 to 124,869. Obviously, a mortality rate of 46% is not observed. Thus, the only explanation that remains is that most countries strongly underestimate the number of affected people. The secondary interest of our model is that it takes into account the rate of change over 3 days, over the past 18 days. Thus it can be used to model the effectiveness of policies to prevent the spread of Covid-19.

## Data Availability

Data are available from:
https://github.com/CSSEGISandData/COVID-19/tree/master/csse_covid_19_data/csse_covid_19_time_series

## ACKOWLEDGMENTS

We thank Matthieu Domenech de Cellès for constructive and supportive advices during the redaction of this manuscript.

## Notes

### Competing Interest Statement

The authors have declared no competing interest.

### Funding Statement

No external funding was received to perform this study.

